# Is death from Covid-19 a multistep process?

**DOI:** 10.1101/2020.06.01.20116608

**Authors:** Neil Pearce, Giovenale Moirano, Milena Maule, Manolis Kogevinas, Xavier Rodo, Deborah A Lawlor, Jan Vandenbroucke, Christina Vandenbroucke-Grauls, Fernando P Polack, Adnan Custovic

**Affiliations:** London School of Hygiene and Tropical Medicine, London, UK; Unit of Cancer Epidemiology, Department of Medical Sciences, University of Turin; ISGlobal, Barcelona, Spain; IMIM (Hospital del Mar Research Institute), Barcelona, Spain; ICREA, Barcelona, Spain; MRC Integrative Epidemiology Unit at the University of Bristol, Bristol, UK; Population Health, Bristol Medical School, University of Bristol, Bristol, UK; Leiden University Medical Center, The Netherlands; Aarhus University Denmark; Department of Medical Microbiology and Infection Prevention, Amsterdam UMC, The Netherlands; Fundacion INFANT, Buenos Aires, Argentina; Department of Pediatrics, Vanderbilt University, Nashville, TN; National Heart & Lung Institute, Imperial College London, United Kingdom

## Abstract

Covid-19 death has a different relationship with age than is the case for other severe respiratory pathogens. The Covid-19 death rate increases exponentially with age, and the main risk factors are age itself, as well as having underlying conditions such as hypertension, diabetes, cardiovascular disease, severe chronic respiratory disease and cancer. Furthermore, the almost complete lack of deaths in children suggests that infection alone is not sufficient to cause death; rather, one must have gone through a number of changes, either as a result of undefined aspects of aging, or as a result of chronic disease. These characteristics of Covid-19 death are consistent with the multistep model of disease, a model which has primarily been used for cancer, and more recently for amyotrophic lateral sclerosis (ALS). We applied the multi-step model to data on Covid-19 case fatality rates (CFRs) from China, South Korea, Italy, Spain and Japan. In all countries we found that a plot of ln (CFR) against ln (age) was approximately linear with a slope of about 5. As a comparison, we also conducted similar analyses for selected other respiratory diseases. SARS showed a similar log-log age-pattern to that of Covid-19, albeit with a lower slope, whereas seasonal and pandemic influenza showed quite different age-patterns. Thus, death from Covid-19 and SARS appears to follow a distinct age-pattern, consistent with a multistep model of disease that in the case of Covid-19 is probably defined by comorbidities and age producing immune-related susceptibility. Identification of these steps would be potentially important for prevention and therapy for SARS-COV-2 infection.

## INTRODUCTION

Respiratory viral infections are a leading cause of mortality globally (1). This has been highlighted by the huge global impact of coronavirus disease 2019 (Covid-19) (2), and the enormous public health threat it poses. Covid-19 is a new and devastating disease, in which the long-term effects of infection are still being discovered. Primary infection with SARS-CoV-2 and a relatively mild disease followed by successful clearance of the virus seems to represent the norm. However, in a minority of patients, there is systematic inflammation and an exaggerated immune response (cytokine storm) resulting in respiratory and multi-organ failure and potentially death (3).

SARS-CoV2 is a particularly aggressive virus for elderly individuals, who represent between 73 and 90% of fatal cases in different regions of the world (4, 5). Mortality, need for intensive care, mechanical ventilation, oxygen requirement, and hospitalizations increase with advancing age in patients with Covid-19 (6–8). Median age among 20,133 hospitalized patients in the UK hospitals with Covid-19 was 73 years, and more men were admitted than women (60% vs. 40%) (9). In hospitalized patients older than 75 years, mortality is 29.4%, and in intensive care units (ICU) can reach a rate of 43.5% (10). It is currently unclear whether children have similar infection rates to adults (11, 12), or are equally infectious; however, when infected they have fewer symptoms, and deaths in children are very rare (13).

Overall, death from Covid-19 increases monotonically and steeply with age. This is in contrast with other respiratory pathogens, such as respiratory syncytial virus (14) and seasonal influenza (15, 16) which cause substantial numbers of deaths in children and have a J-shaped pattern with age. In contrast, pandemic influenza viruses often cause severe disease in middle-aged adults (17–19), even without pre-existing co-morbidities (20), suggesting different pathogenesis from that for seasonal influenza. One exception was mortality from pandemic influenza in 2009, which had different age-distribution to other influenza pandemics (15); severe disease was relatively high in young adults, and appeared to be driven by a non-protective anamnestic response by previously acquired antibodies to the new virus. Thus, there are some notable features of Covid-19 death which suggest that it has a markedly different relationship with age than is the case for most other severe respiratory pathogens.

The mechanisms explaining excess Covid-19 mortality and ICU admissions in elderly are unknown (21). Having an underlying condition such as hypertension, diabetes, cardiovascular disease, severe chronic respiratory disease and cancer increases the risk (22, 23) – e.g. an Italian study(22) found that 62% of cases and 85% of Covid-19 deaths had at least one comorbidity, whereas a similar study in China(23) found estimates of 25% and 70% respectively. However, a strong predictor of mortality after adjusting for major comorbidities is increasing age itself (9). Furthermore, the extreme rarity of deaths in children suggests that infection alone is not sufficient to cause death; rather, one must have gone through a number of changes, either as a result of unknown aspects of aging, or as a result of chronic disease(effectively speeding up the aging process), increasing susceptibility, leading to severe disease and death.

These characteristics of Covid-19 death are consistent with the multi-step model of disease, a model which has primarily been used for cancer (24, 25), and more recently for amyotrophic lateral sclerosis (ALS) (26). Briefly, if one assumes that a disease outcome occurs due to the accumulation of a number of discrete steps throughout life, then the incidence of the outcome (this could be the incidence of disease occurrence, or in the case of Covid-19 the incidence of death) will be proportional to age to the power of n-1, where n is the total number of steps involved. Thus, a plot of log (incidence) against log (age) will be a straight line, and the slope (n-1) will be one less than the number of steps (n).

For epithelial tumours, the number of steps appears to be about 6 (this is a population average since some subtypes may involve different numbers of steps), and in some instances (e.g. colorectal cancer), the relevant steps have been identified (25). Similar findings have been obtained for ALS, whereas some other chronic neurological conditions (e.g. multiple sclerosis) do not follow a multistep pattern (26). It thus appears that some diseases are ‘digital’ and involve discrete steps which produces the log-log linear relationship with age, whereas others are analogic and do not show these same age patterns.

Given what is known about the age-distribution of Covid-19 death, and its clinical characteristics, we considered that it would be useful to explore where Covid-19 death followed an age-pattern consistent with the multistep model. We were also interested in comparing the findings in males and females, since males have a higher death rate in each age-group (9, 27). The mechanisms explaining excess Covid-19 mortality in males are unknown (21), and it has been suggested that this may be from an inherent characteristic of being male (28). In multistep model terminology, this would mean that males were born with one step already in place, analogous to being born with a particular genetic mutation which accounts for one step of a multistep cancer process. In this scenario, males would have higher death rates at each age-group, but their overall age slope would be one lower (e.g. 4 instead of 5), since one step had already been acquired at birth.

## METHODS

### Issues with available data for testing the multistep model for Covid-19 deaths

As we have discussed elsewhere (29), there are a number of potential problems with reported Covid-19 infection and mortality data. These include non-random population sampling for testing (mostly symptomatic people get tested) which means that diagnosed cases are likely to be more severe, misclassification of infection and/or misclassification of cause of death. Moreover, there is little valid information on the underlying population infection rates. This means that estimates are available for the case fatality rate (CFR = deaths/diagnosed cases), but few estimates are available for the infection fatality rate (IFR = deaths/total infected). For the current analyses, using IFRs may have been preferable, but in almost all cases only CFRs were available in our data sources. Even for the CFR, there are significant problems in estimating it accurately (30, 31), as is the case for pandemic influenza (32). Under-ascertainment of infections is most likely to be a problem in younger age-groups where there is likely to be less testing, because there are fewer symptomatic infections (13).

### Data sources

We needed data on Covid-19 CFRs or IFRs by age-group, ideally separately for males and females. We sought potential data sources through literature searches, examination of national statistics online, and through enquiries with colleagues in different countries. In some instances, population death rates (i.e. with total population denominators) were available, but these are not appropriate for the multistep model in the current analysis which focusses on case fatality. Therefore, we restricted the analysis to data with CFRs (cases as denominators) or IFRs (infections as denominators).

#### China

The data for estimates of the CFR and IFR for China were taken from the publication by Verity et al (33). Briefly, these involved 44,672 PCR-confirmed cases in Wuhan and elsewhere in China during the period 1^st^ January to 11^th^ February 2020, extracted from the WHO-China Joint Mission Report (34); during the same period 1,023 Covid-related deaths were reported across China. The main analyses in the publication were adjusted for censoring, demography and under-ascertainment. We report the findings using the unadjusted estimates, since these are more comparable to the data from other countries. However, only the adjusted IFR is available in the report and therefore to compare the findings for the CFR with those for the IFR in the China data we used the adjusted estimates for both.

#### South Korea

Data were retrieved from the online website of the Korean Centre for Disease Control (KCDC). KCDC included all confirmed Covid-19 cases collected by the surveillance system from 20^th^ January to 17^th^ of March 2020. During the identified timespan a total of 8,320 cases and 81 Covid-19 related deaths had been confirmed in South Korea (35).

#### Japan

The data for Japan were obtained from the website of the Ministry of Health, Welfare and Labour, 6^th^ May 2020; up to that date there were 15,300 cases and 537 deaths reported.

#### Italy

The data for estimates of the CFR for Italy were obtained from the scientific report released by the National Health Institute (ISS) on April 17^th^ (36). The data refers to all laboratory-confirmed cases collected by the national surveillance system from 21^st^ February to 16^th^ April 2020. During the identified period 159,107 Covid-19 cases and 19,996 Covid-related deaths were reported in Italy.

#### Spain

The data was obtained from the official reports of the Spanish government (Ministry of Health and the Instituto de Salud Carlos III). It includes the reported cases, recovered and deaths by age category (37). During the reporting period up till 18^th^ May 2020, there were 239,125 Covid-19 cases and 19,186 deaths.

As a comparison, we also obtained similar data for three other respiratory diseases (seasonal influenza, pandemic influenza and Severe acute respiratory syndrome-SARS). The details of the data sources, and findings for these, are given in the Supplementary Appendix.

### Data analysis

Under-ascertainment of cases is mostly likely to be a problem in younger age-groups, and deaths are rare in this age-group and the rates are therefore unstable (ln (CFR) cannot be calculated when there are zero deaths). We therefore included all ages (0–99 years) in the descriptive analyses, but restricted the multistep analyses to ages 30 years and over (an approach that has been used in some other analyses of Covid-19 mortality (38)). We analysed age-specific (and where available, sex-specific) death rates (i.e. CFRs and IFRs) in ten-year age-groups (for publications which reported five-year age-groups we amalgamated these into ten-year groups in order to standardize the analysis). We used the mid-point of each age-group and regressed the natural log of the CFR or IFR against the natural log of age, using standard linear least-squares regression. As in similar previous analyses (26), we used unweighted regression so that the estimated slope was not biased towards the older age-groups where there were many more deaths. The multistep model predicts a slight flattening of the curve in the youngest and oldest age-groups - this occurs because deaths in young age-groups often involve people who have inherited at least one step, and therefore have a high risk but a lower slope – in the oldest age-groups many people will already have accumulated 1–2 steps, so once again the risk is higher but the slope is lower (25). For the multistep model, we are interested in the overall slope with equal weight on all of the age-groups (essentially it is the middle age-groups which are predicted to provide the slope which is one less than the number of steps). We did the regressions for each country, by sex (data available for Spain and Italy), and we also did combined analyses adjusting for country; these were weighted by the total number of cases in each country’s dataset.

## RESULTS

### Multi-step analyses for Covid-19

For all analyses, we found a linear log-log relationship between the CFR and age, with a small flattening of the curve in the youngest and oldest age-groups (Table 1). All five countries had an overall slope of about 5. Figures 1 and 2 show the findings for the CFR rate by age, and the plot of ln (CFR) against ln (age), for all five countries combined. The country-specific findings are shown in Figures S1-S5 (Supplementary Appendix).

**Table 1:** Slope of ln (death rate) versus ln (age) for the five countries, for CFR, for the total population and separately for females and males where available.

| **Country** | **N of cases** | **Total** | **95% CI** | **Females** | **95% CI** | **Males** | **95% CI** |
| --- | --- | --- | --- | --- | --- | --- | --- |
| China | 44,672 | 4.88 | 3.96-5.80 |  |  |  |  |
| South Korea | 8,320 | 5.66 | 3.01-8.31 |  |  |  |  |
| Japan | 15,300 | 5.77 | 4.75-6.79 |  |  |  |  |
| Spain | 239,125 | 4.94 | 3.95-5.93 | 5.37 | 4.25-6.50 | 4.63 | 3.80-5.46 |
| Italy | 159,107 | 4.93 | 3.70-6.15 | 5.43 | 4.00-6.86 | 4.76 | 3.90-5.61 |
| **Total** | **466,524** | **4.94** | **4.51-5.37** | **5.40** | **4.67-6.12** | **4.68** | **4.19-5.17** |

**Figure 1.**
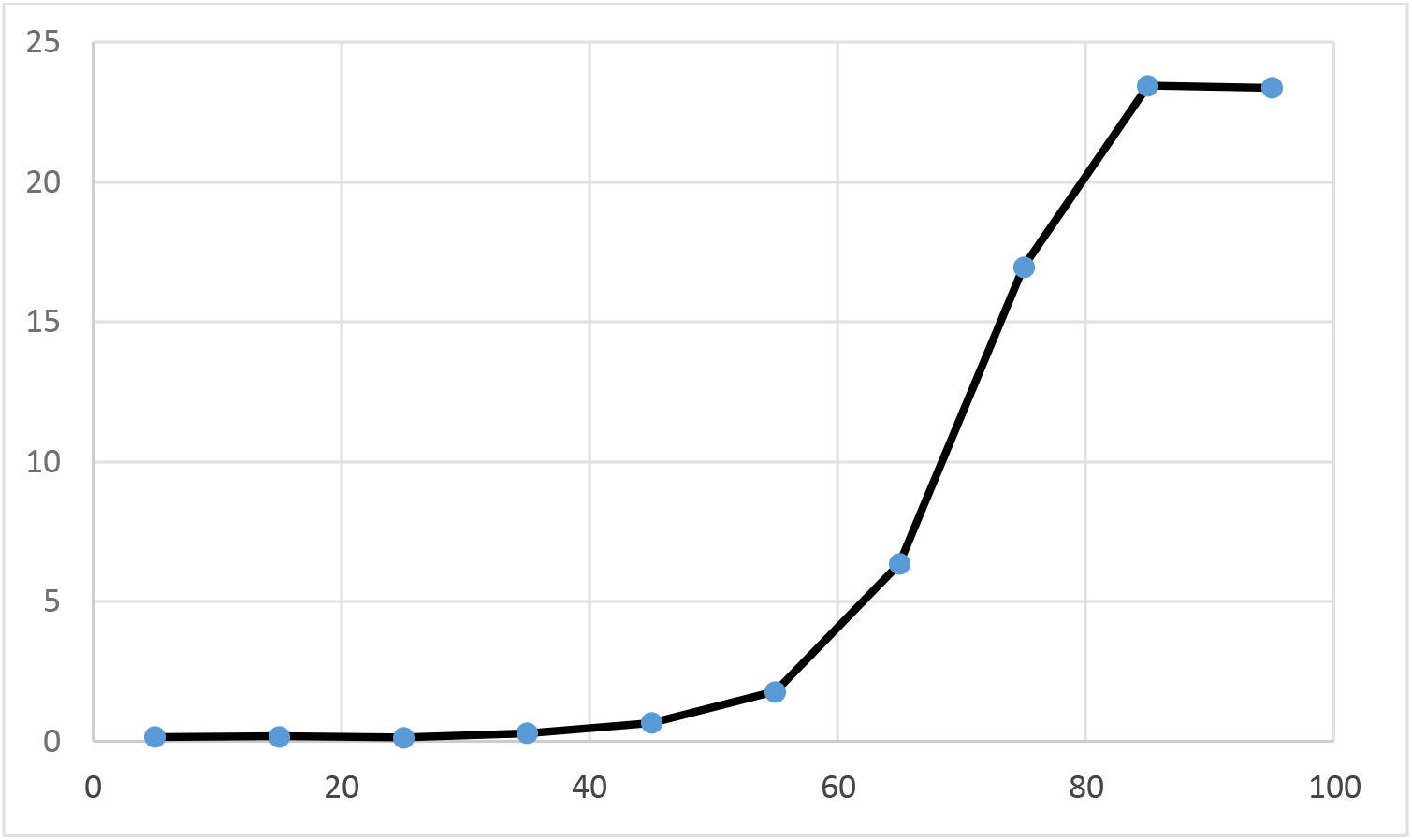
CFR by age for the four countries combined.

**Figure 2.**
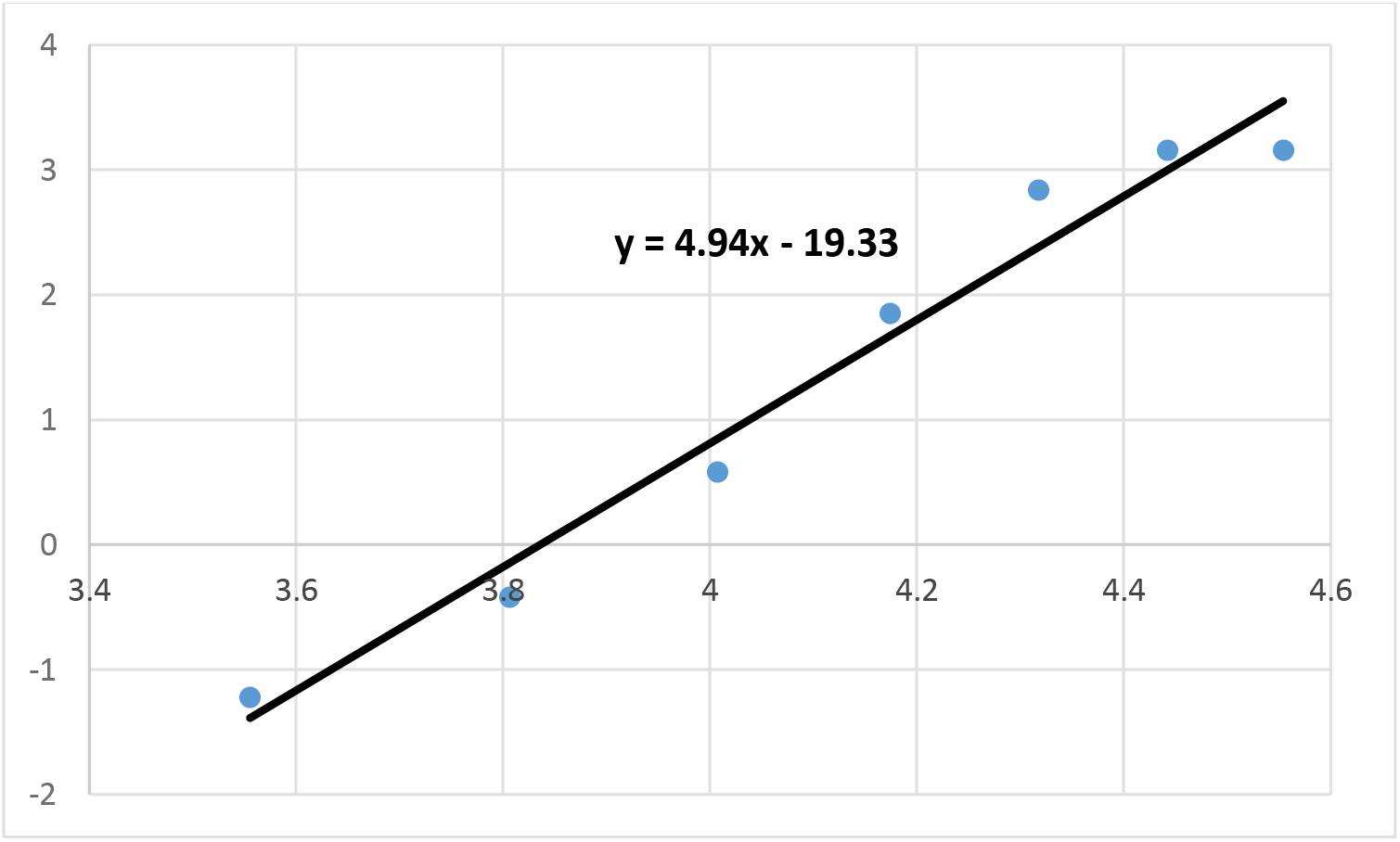
Ln (CFR) versus ln (age) for the four countries combined.

For the two countries where we had data separately for males and females (Italy and Spain), the male death rates were higher at each age-group, but the slope was lower - by 0.7 (Figures 3 and 4).

**Figure 3.**
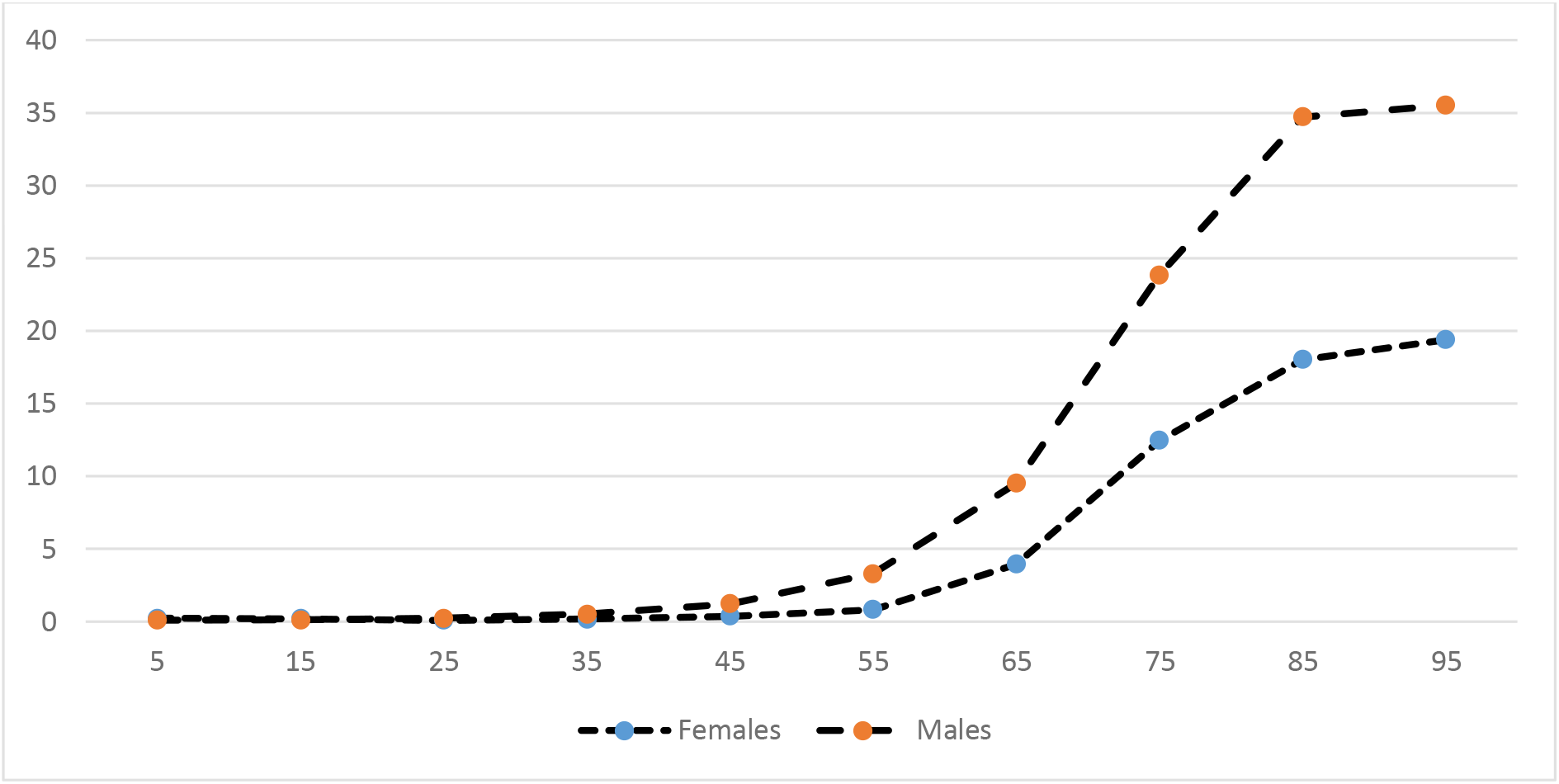
CFR by age for two countries combined (Italy and Spain), by sex.

**Figure 4.**
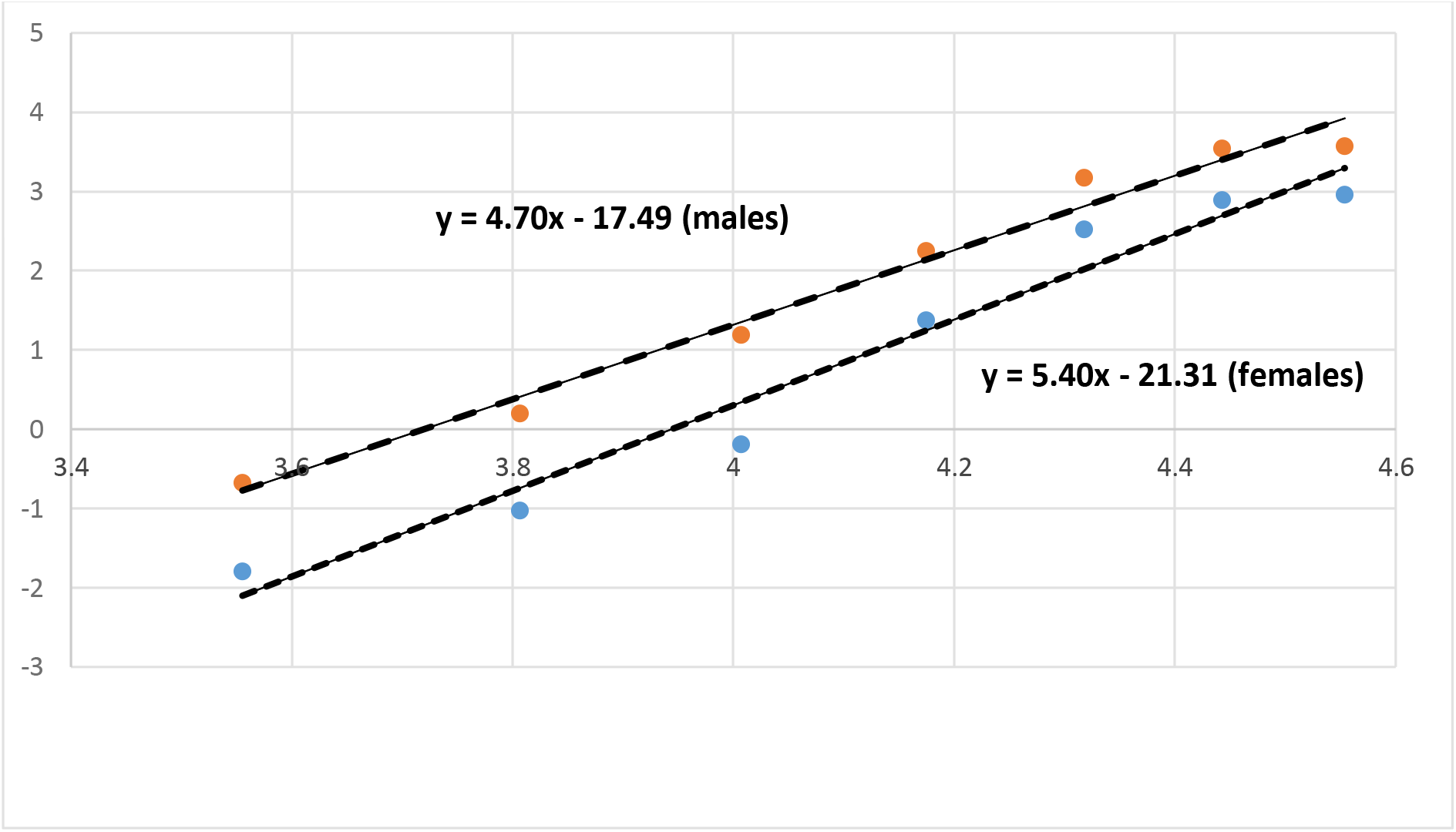
Ln (CFR) versus ln (age) for two countries combined (Italy and Spain), by sex.

In one country, China, we had data for both CFR and IFR, and these yielded very similar findings (not shown in table); these used adjusted data (see above), and yielded slopes of 5.5 for the CFR and 5.4 for the IFR (Figures S6 and S7, Supplementary Appendix).

### Multi-step analyses for other respiratory virus diseases

The findings for selected other respiratory virus diseases (Seasonal influenza, Pandemic influenza and SARS) are shown in Figures S8-S10 (Supplementary Appendix). Seasonal influenza (Figure S8) and pandemic influenza (Figure S9) showed age-patterns that were different from each other, and markedly different from that for Covid-19 (Figure 2). Although seasonal influenza generally showed an exponential increase with age, there were also high death rates in the 0–4 age-group (i.e. the overall pattern is J-shaped). Data on mortality in hospitalised influenza cases in England from January 2000 to August 2007 (16), i.e. during non-pandemic years, did not clearly show a log-log pattern, and the fitted slope is only 1.4 (Figure S8). Pandemic influenza (H1N1) showed a very different age-pattern, with peak mortality in adulthood. Modelling data from England (39) on mortality due to 2009 H1N1 influenza pandemic shows a J-shaped pattern mortality in the youngest age-group, with no log-log pattern, and the fitted slope is close to zero (Figure S9). SARS showed a roughly linear log-log relationship (Figure S10), but with a lower slope (3.6) than was observed for Covid-19 (Figure 2). Thus, SARS showed a similar log-log age-pattern to that of Covid-19, albeit with a lower slope; in contrast, seasonal and pandemic influenza showed quite different age-patterns, with little evidence of a log-log relationship, and with substantially lower slopes.

## DISCUSSION

As hypothesized, we found a linear log-log relationship between the CFR and age, with a small flattening of the curve in the youngest and oldest age-groups (as predicted by the multistep model (25)). All five countries gave similar values for the overall slope (Table 1), i.e. about 5, indicating a multistep process of about 6 steps. As with other applications of the multistep model (e.g. cancer), this is a population average, and it may well be that some pathways to the outcome may involve a different number of steps(25).

A number of limitations of the data should be recognized.

Firstly, the appropriate denominator depends on what the hypothesis is. For cancer, where the multistep model has been most commonly used, the hypothesis is that there are steps involved in developing cancer, so the outcome/numerator is incidence and the denominator is total population; most risk factors for cancer that follow the multistep model (e.g. a genetic mutation) affect incidence and usually not survival (25). In the current context, the outcome is Covid-19 death, and the hypothesis relates to mortality in those who become infected. However, in most cases, only case fatality rates (CFRs) were available, and the denominator was diagnosed cases rather than all infections. A related problem is selective case identification - most cases are identified by testing symptomatic people, and non-symptomatic cases are largely missed, and even in those tested there will be false positive and false negative results (this is also a problem with other respiratory diseases such as pandemic influenza (32)). Given all of these uncertainties, the available data that we have used for these analyses is likely to be subject to inaccuracy and misclassification (of outcome, denominator, or both). However, these uncertainties will not bias the analysis unless they operate with different strengths at different age-groups. For example, if the problem is just that the CRF is double the IFR at each age-group greater than 30 years (because at each age-group, only one-half of the infections are tested and diagnosed), this will affect the absolute value of the CFR but will not affect the slope of the log-log linear trend. Furthermore, it is noteworthy that we have obtained very similar findings for the CFR in several different populations and contexts, with differing methods for ascertaining cases.

If we can assume that the findings presented here are valid, given all the uncertainties of the data, what do they mean? The multistep model has been used for cancer since 1954, but with a few exceptions (e.g. colorectal cancer (25)) the relevant stages have not yet been identified. Those that have been identified typically involve mutations in DNA or other cellular changes which lead to cell proliferation. The model has produced a number of testable hypotheses, e.g. relating to the stage (step) at which various carcinogens act (40), the dose-response relationships for factors such as smoking, and the changes in the age-incidence patterns after smoking cessation (25). The application of the multistep model to ALS (26) is beginning to yield similar benefits, including clarifying the role of genetic susceptibility which increases the disease rate at each age-group but produces a lower slope compared with persons without the genetic susceptibility (41). Thus, identifying diseases that show a multistep pattern provides the foundations for identifying the steps and being able to intervene across the life-course. Importantly, for Covid-19 death if a multistep pattern is identified, it illustrates that identification of final (trigger) steps in (elderly) patients could prevent severe disease and death.

We also conducted similar analyses for selected other respiratory diseases. SARS showed a similar log-log age-pattern to that of Covid-19, albeit with a lower slope (indicating a smaller number of steps); in contrast, seasonal and pandemic influenza showed quite different age-patterns. Thus, death from Covid-19 and SARS appears to follow a distinct age-pattern, consistent with a multistep model of disease.

Given that the stages involved in multistep cancer causation have mostly not yet been identified, and that SARS-Cov-2 is a newly discovered virus, it is not surprising that the stages involved in Covid-19 mortality are not currently readily identifiable. However, they clearly involve changes associated with chronic conditions such as hypertension, diabetes, cardiovascular disease, severe chronic respiratory disease and cancer. They also involve other cumulative changes associated with aging (e.g. one change may be the nasal expression of ACE-2 receptors with aging (42)), since age itself is a risk factor for Covid-19 death. Collectively, these comorbidities, and age itself, appear to be markers of immune-related susceptibility, which makes the difference between experiencing a mild illness, or a severe illness possibly resulting in death.

Given the rapidity of Covid-19 death following infection, it is likely that SARS-Cov-2 itself acts at a late stage of the multistep process, with the previous stages having occurred before infection, throughout the life-course. Simplistically, if someone has already accumulated a number of the necessary steps (from having one of these chronic conditions, or from other cumulative changes associated with aging), then they are ‘primed’ to have a serious case of Covid-19 if and when infection occurs. If they are not so ‘primed’, then infection can still occur, but is usually mild. This is most obvious for infections in children - who have not had one of these chronic conditions (and are not old) – where the infection is almost always mild.

However, it is not so clear what these steps may be. Does having diabetes or a high BMI, for example, involve a specific biological change (analogous to those observed for cancer and ALS) which is an intrinsic part of the process leading to Covid-19 mortality? If this is the case, are these conditions clearly-defined ‘steps’ which can potentially be identified? If so, one would expect that people who have one of these conditions (e.g. diabetes), would have a higher CFR for Covid-19 death at each age-group, but a lower slope – this is a hypothesis that can be tested in future as more data with sufficient numbers of the underlying conditions emerge. On the other hand, are these conditions, like age, just markers of more general increases in immune-related susceptibility which accumulate across the life-course? In which case there must be other steps that ‘prime’ someone for severe disease when they are infected. Whether or not distinct risk factors can be identified for the underlying steps, the clear log-log patterns of the CFR rate with age that we have identified indicate that there is a potential to identify population subgroups which are ‘primed’ for severe disease if they experience a SARS-CoV-2 infection. It is also striking that we see a similar pattern with SARS (another Coronavirus), perhaps indicating a similar mechanism for mortality, whereas we see different age-patterns for other severe respiratory infections, perhaps indicating that different mechanisms are operating.

We found that the male CFR was higher than that in females at each age-group, but that the slope of the regression was 0.7 lower in males. This is close to the difference of 1.0 which was the a priori hypothesis. A difference of 1 would suggest that males are born with one step in place. We propose that this step may be related to a relative deficiency in innate immune responses to viruses in males compared to females. Recent analysis in a population-based birth cohort has shown that interferon (IFN)-α, –β and –γ responses to three common respiratory viruses and two viral mimics are deficient in males compared to females, indicating that the excess Covid-19 mortality in males is likely at least in part explained by impaired innate anti-viral immune responses in males compared to females (Prof S Johnston, personal communication). Endosomal-expressed Toll-like receptors (TLRs) 7/8/9 are engaged by positive strand RNA viruses such SARS-CoV-2, which require endosomal processing as part of viral entry into cells(43). Their activation results in production of type I IFNs, which is an important step for the induction of antiviral immunity. TLRs 7 and 8 are encoded by loci on X chromosome locus, and biallelic expression of X-linked genes could enhance TLR7-8 expression in female immune cells (44), thereby providing a mechanistic explanation for our observation. We propose that the notion that biallelic expression of X-linked TLR7-8 is unlikely to be strictly binary, and will not occur in every female, could explain 0.7 (rather than 1) difference.

Our findings should be regarded as preliminary, and require further replication in other populations. Moreover, their etiological significance is not yet clear, though as described above specific hypotheses, such as whether diabetes is likely one of the steps, can be tested as more data emerges. Nevertheless, the patterns are strikingly consistent across the countries studied. These findings are consistent with a multistep model of disease involving a six-step process that in the case of SARS-COV-2 is probably defined by comorbidities and age producing immune-related susceptibility. Identification of these steps would be potentially important for prevention and therapy for SARS-COV-2 infection.

## Data Availability

This manuscript involves re-analyses of published data.

## Acknowledgements

We thank Elizabeth Brickley, Stephen Evans, Matthew Fox and Judith Glynn for their comments on the draft manuscript. We also thank Yasuyuki Gondo for supplying the data from Japan.

